# Effect of Behaviourally Informed Text Messages to Promote Retention in HIV Care: A Randomized Trial in Ekurhuleni District, South Africa

**DOI:** 10.1101/2023.10.29.23297725

**Authors:** Caroline Govathson, Sophie Pascoe, Candice Chetty-Makkan, Laura Schmucker, Preethi Mistri, Harsha Thirumurthy, Tonderai Mabuto

## Abstract

**Introduction:** Sustained engagement in care among people living with HIV is vital for realising the individual and public health benefits of antiretroviral therapy (ART). We examined whether mobile phone-based text messages that leveraged behavioural science principles promoted timely clinic attendance among ART recipients.

**Methods:** We conducted a randomized controlled trial in three primary health clinics in Gauteng Province, South Africa between July 2021 and December 2021. ART recipients with an upcoming clinic appointment were randomised to standard of care appointment reminders or three enhanced reminders that leveraged behavioural science principles of loss aversion, social norms, and altruism. The primary outcome was a timely clinic visit, on or before the scheduled appointment day. Poisson regression with clinic fixed effects and controls for age, sex, and ART duration was used to examine the effect of the enhanced reminders on the primary outcome.

**Results:** Among 1539 participants, 690 (44.8%) were male and median time on ART was 11 months (IQR, 3.7-51.9). The primary outcome of timely clinic visit was 50.3% in the standard of care arm, and similar in the loss aversion arm (53.5%; adjusted risk ratio, ARR 1.1; 95% CI: 0.9-1.2), social norms arm (48.0%; ARR 1.1; 95% CI: 0.8-1.1), and altruism arm (50.9% ARR 1.0; 95% CI: 0.9-1.5). In subgroup analyses, messages framed with loss aversion increased the timely clinic visits among participants with ART duration <90 days (ARR=1.37; 95% CI: 1.08-1.74).

**Conclusions:** The use of behavioural science principles to frame reminder messages did not increase timely clinic visits among HIV care recipients. Among those who recently initiated ART, however, loss aversion framing increased timely clinic visits. Future research should explore alternative behavioural science principles to revise health communication with HIV care recipients.

**Clinical Trials Number:** NCT05010291

## Introduction

In recent years, an increasing number of studies have shown how shifts in the needs, values and preferences of many people on antiretroviral therapy (ART) often translates to cyclical patterns of clinical care, characterised by varying periods of disengagement, temporary lapses in ART, and reengagement (1,2). These cyclical patterns are often associated with suboptimal viral suppression and increased risk of onward transmission of HIV (1,3,4).

With many countries implementing same-day ART initiation, the challenges with interruption in care and retention are more pronounced in the first six months after ART initiation (5,6). Clinic appointment attendance has emerged as a strong predictor for longer term retention of ART clients since missed appointments can lead to a reluctance to return to care and ultimately result in disengagement from HIV treatment (7–9). Beyond the risk of disengagement from ART, when care recipients are late for, or miss their scheduled appointments, this may place additional pressure on already constrained clinic staff to trace and bring clients back to care, negatively affecting the quality-of-service delivery (9–11).

Healthcare systems need be highly agile in developing new strategies or enhancing existing ones to promote continued engagement in HIV care and adherence to ART among recipients of care. For many years, text message appointment reminders have been used to promote timely and continuous engagement of clients receiving chronic care (12–14). In the early years of scaling up text message appointment reminders, these strategies were mostly targeted at addressing clients’ forgetfulness and confusion over appointment dates (15,16). However, more recently, studies have begun to explore how text message appointment reminders may minimise lapses in ART by influencing other key decision-making points that influence and individual’s pattern of clinical care engagement. Notably, many of these studies build on emerging evidence from behavioural science research that shows how individual-level decision making can be influenced by minor changes in how information is presented – commonly referred to as the framing effect.

Although there is mixed evidence on how well text message appointment reminders improve client outcomes across different stages of the HIV care continuum (17), their low-cost, high acceptability, and high reach to recipients of care, justifies the need to explore whether the content of text messages appointment reminders can contribute to better client outcomes. This is particularly important for South Africa, a low-middle income country that provides ART to about 5.7 million people (18), many of whom exhibit cyclical patterns of care engagement (1,19). One study assessing testing and restarts reported 71% of individuals retesting had previously started ART (1).

In a randomised control trial in South Africa, we sought to evaluate whether framing effects on text message appointment reminders, informed by behavioural science, would effectively promote adherence to scheduled clinic appointment among ART recipients.

## Methods

### Study Design and Setting

This study was an individually randomized controlled trial conducted in three public sector clinics in Ekurhuleni District, Gauteng Province, South Africa. The three clinics were purposively selected based on their large number of ART recipients, routine use of text message appointment reminders and a high burden of missed appointments among care recipients (18). Ekurhuleni District has the second-highest district-level HIV prevalence in South Africa (14.3%), and high levels of unemployment and poverty, increase the occurrence of cyclical care engagement patterns among ART recipients.

### Participants

Individuals living with HIV who were ≥18 years old, receiving ART or about to initiate ART were eligible for the study. Care recipients without cell phone contact details, those who had self-transferred out of the study clinic, or were known to be deceased were excluded. A waiver of participant consent was obtained from the University of Witwatersrand Human Research Ethics Committee, the Ekurhuleni Research Ethics Committee, and the University of Pennsylvania Institutional Review Board, as the study did not alter routine care delivery, did not involve more than minimal risk to recipients of care, and could not have been practicably carried out without a waiver of client consent.

Block randomisation sequences (block size 8) were generated by an independent biostatistician, separate from the study implementation team at the study clinics. Participants were randomly assigned in a 1:1:1:1 ratio to either the standard of care text message arm or three text message appointment reminders enhanced with framing effects (described in the next section). Separate randomisation sequences were created for each study clinic. While the study arm allocations were unmasked for randomisation assignment by routine healthcare providers who were involved in the study at each clinic, all study staff performing outcome assessments were masked to the study assignments until all outcome data were collected.

### Intervention Design

The study had four arms, with three behavioural science-informed text message arms and one standard-of-care (SOC) text message arm. The content and framing of the text message appointment reminders in the intervention arms were developed by a panel of behavioural science experts and reviewed by routine healthcare providers designated to assist with care navigation and sending appointment reminders (hereafter referred to as case officers) at the study sites to ensure accuracy and ease of understanding. All messages were developed in English and translated into two local languages (Sesotho and isiZulu) commonly used in the study setting.

The SOC message contained the name of the clinic sending the message, information about clinic operating hours, reminded them of their appointment, and prompted them to attend the visit. The text messages in the three intervention arms used framing effects to modify the SOC message regarding how care recipients were reminded of their appointment and how they were prompted to attend the visit.

The text message in the first intervention arm was framed around the concept of loss aversion which is grounded in the human tendency to prefer avoiding losses over receiving an equivalent gain. The loss aversion message sought to nudge care recipients that an appointment day was “reserved for them” and encouraged them to not lose the opportunity to see a provider during the reserved day.

The text message in the second intervention arm was framed around the concept of social norms, which relates to how people engage in social comparison and are strongly influenced by others’ behaviours. The social norms message sought to nudge care recipients that on-time clinic attendance was the norm for most individuals who were staying on track and maintaining good health.

The text message in the third intervention arm was framed around the concept of altruism, which focuses on sacrifices made to benefit others. The altruism framing message highlighted the importance of going to the clinic visit on time to protect one’s health and the health of people they love. Participants received only one text message appointment reminder during the study period.

### Data Collection

Case officers used a web-based text messaging service to send one-way text message appointment reminders to each participant one week (seven days) before the participant’s scheduled appointment. The web-based service also indicated whether each message was successfully delivered to the participant. Case officers abstracted outcome information from care recipient clinic paper folders and electronic health records.

### Data analysis

The primary outcome was a timely clinic visit, defined as having occurred if the participant went to the clinic on or before the scheduled date of their clinic appointment. Participant baseline characteristics were described using proportions for categorical variables and either medians and interquartile ranges (IQRs) or means and standard deviations for continuous variables. Poisson regression models was used to estimate the effect of each intervention. In adjusted analyses, we controlled for participant age, gender, ART duration at the time of the scheduled clinic appointment, and whether successful message delivery occurred (based on text message system reports).We conducted an intention to treat analysis, including participants whose messages were not delivered. The same analyses were performed for four different participant subgroups based on antiretroviral therapy duration (<90, 90-180, 180-365, >365 days). This subgroup analysis was not pre-specified, but the intervention effect was examined in each subgroup given the variation in antiretroviral therapy adherence and appointment attendance patterns among people living with HIV who have been on antiretroviral therapy for different durations. The study was approved by the research ethics committees at the University of the Witwatersrand # 210308 and University of Pennsylvania # 849826.

## Results

Between July 2021 and October 2021, 1635 ART recipients had a scheduled clinic appointment at the three study specific clinics. We excluded 10 individuals who were aged <18 years, 31 individuals who did not have available phone numbers, 53 who had duplicate participant identification numbers, and 2 individuals for whom the purpose of their clinic visit was unknown **(Figure 1)**.

**Figure 1.**
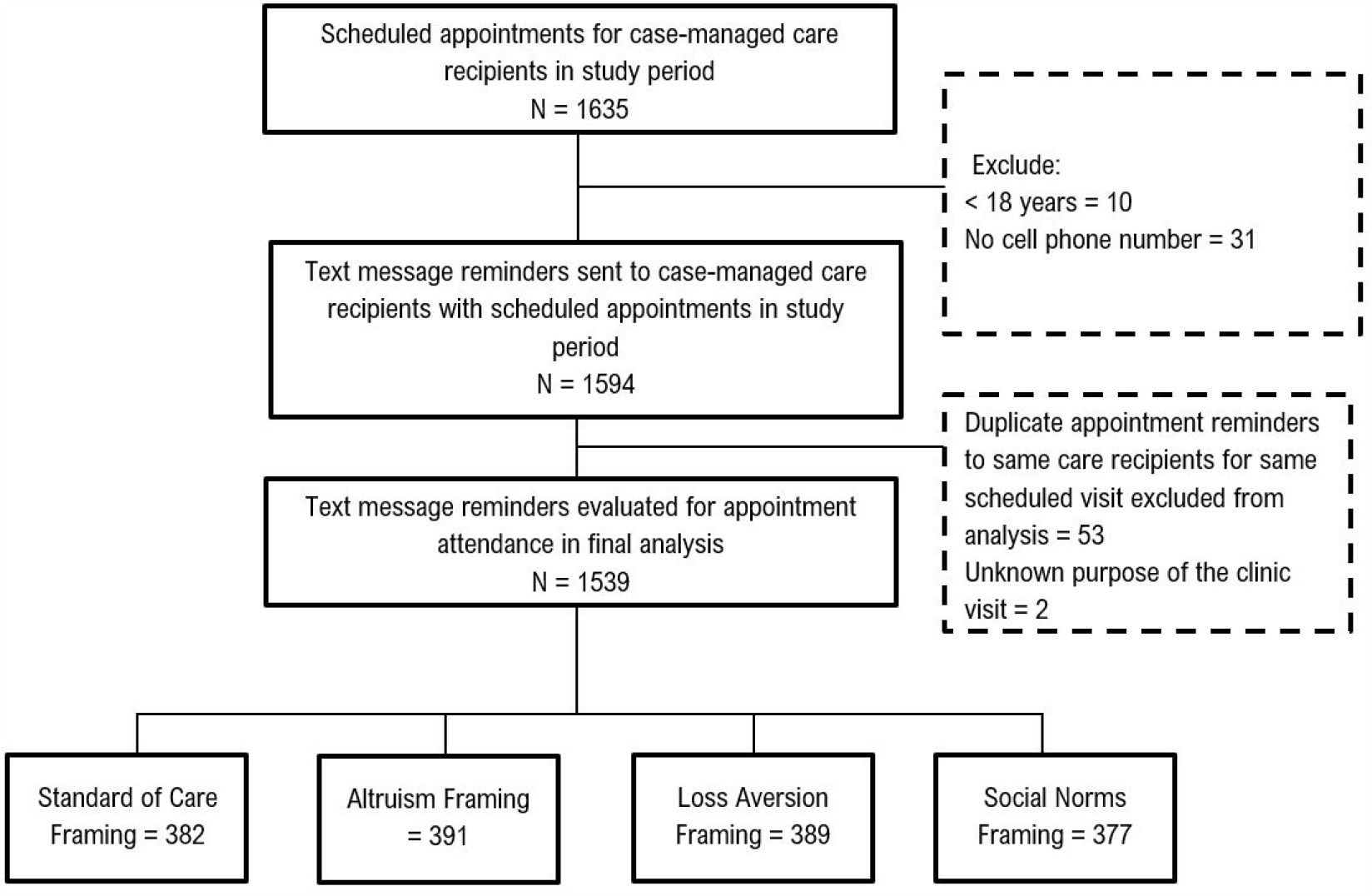
Flow of procedures describing assignment of the groups receiving the text messages between July-October 2021 from three clinics in Ekurhuleni, South Africa

For the 1539 care recipients enrolled in the study, 55.2% were female and the median age was 37 years (IQR, 31-43). Median time on ART was 11.6 months (IQR 3.8-52.1 months) and the purpose of the clinic visit for most (80.7%) participants was ART refill. Baseline characteristics including age and sex were similar among the four intervention groups and no significant differences were found (**Table 1)**. Around 60% of participants did not receive the text messages that were sent to them. Delivery rates did not vary by study arm.

**Table 1:** Baseline characteristics of study participants.

|  | Variable | Total<br>(n=1539) | SOC<br>(n=382) | Loss Aversion<br>(n=389) | Social Norms<br>(n=377) | Altruism<br>(n=391) |
| --- | --- | --- | --- | --- | --- | --- |
| **Clinic** | Clinic 1 | 970 (63.0%) | 242 (63.4%) | 242 (62.2%) | 239 (63.4%) | 247 (63.2%) |
|  | Clinic 2 | 255 (16.6%) | 62 (16.2%) | 67 (17.2%) | 61 (16.2%) | 65 (16.6%) |
|  | Clinic 3 | 314 (20.4%) | 78 (20.4%) | 80 (20.6%) | 77 (20.4%) | 79 (20.2%) |
| **Sex** | Male | 690 (44.8%) | 160 (41.9%) | 173 (44.5%) | 164 (43.5%) | 193 (49.4%) |
| **Age** | 18-29 | 310 (20.1%) | 81 (21.2%) | 74 (19.0%) | 78 (20.7%) | 77 (19.7%) |
|  | 30-39 | 644 (41.9%) | 145 (38.0%) | 171 (44.0%) | 154 (40.9%) | 174 (44.5%) |
|  | 40-49 | 432 (28.1%) | 118 (30.9%) | 110 (28.3%) | 106 (28.1%) | 98 (25.1%) |
|  | ≥ 50 | 153 (9.9%) | 38 (10.0%) | 34 (8.7%) | 39 (10.3%) | 42 (10.7%) |
| **Duration on ART** | < 90 days | 265 (17.2%) | 67 (17.5%) | 61 (15.7%) | 68 (18.0%) | 69 (17.7%) |
|  | 90-181 days | 234 (15.2%) | 53 (13.9%) | 58 (14.9%) | 65 (17.2%) | 58 (14.8%) |
|  | 182-364 days | 279 (18.1%) | 67 (17.5%) | 75 (19.3%) | 62 (16.5%) | 75 (19.2%) |
|  | ≥ 365 days | 756 (49.1%) | 193 (50.5%) | 195 (50.1%) | 181 (48.0%) | 187 (47.8%) |
|  | Missing | 5 (0.3%) | 2 (0.5%) | 0 (0.0%) | 1 (0.3%) | 2 (0.5%) |
| **Purpose of appointment** | ART initiation | 64 (4.2%) | 16 (4.2%) | 14 (3.6%) | 15 (4.0%) | 19 (4.9%) |
|  | ART refill | 1242 (80.7%) | 305 (79.9%) | 310 (79.7%) | 303 (80.4%) | 324 (82.9%) |
|  | Specimen | 233 (15.1%) | 61 (16.0%) | 65 (16.7%) | 59 (15.7%) | 48 (12.3%) |
| **\*Time of month message sent** | Month end | 597 (38.8%) | 147 (38.5%) | 158 (40.6%) | 142 (37.7%) | 150 (38.4%) |
| **\*\*Received reminder call** | Yes | 1035 (67.3%) | 252 (66.0%) | 272 (69.9%) | 254 (67.4%) | 257 (65.7%) |
| **Message delivered successfully** | Delivered | 921 (59.8%) | 228 (59.7%) | 231 (59.4%) | 224 (59.4%) | 238 (60.9%) |

\* month end was defined as message sent between 25<sup>th</sup> of a month and the 7<sup>th</sup> of the following month
\*\* Some participants successfully received a phone call as a reminder as part of routine follow-up

The primary outcome of timely clinic visits was similar across study arms, with no significant difference due to the text messages in either unadjusted models or adjusted model. In the SOC arm, 50.3% of participants had a timely clinic visit. Compared to the SOC arms, timely clinic visits were similar in the loss aversion arm (53.5%; adjusted risk ratio, ARR 1.06; 95% CI: 0.93-1.22), social norms arm (48.0%; ARR 0.94: 95% CI: 0.83-1.08), and altruism arm (50.9%; ARR 1.01; 95% CI: 0.89-1.15). Participants receiving ART for <90 days were more likely to have timely clinic visits (ARR 1.25; CI: 1.08-1.43) (**Table 2)**.

**Table 2.** Effect of different text messages reminders on timely clinic visits.

|  | **SOC** | **Loss Aversion** | **Social Norms** | **Altruism** |
| --- | --- | --- | --- | --- |
| **All participants (n)** | 382 | 389 | 377 | 391 |
| Attended clinic appointment, N (%) | 192 (50.3%) | 208 (53.5%) | 181 (48.0%) | 199 (50.9%) |
| Unadjusted risk ratio (95% CI) | Ref | 1.06 (0.93 - 1.22) | 0.95 (0.83 – 1.10) | 1.01 (0.88 – 1.16) |
| Adjusted risk ratio* (95% CI) |  | 1.06 (0.94 – 1.20) | 0.94 (0.83 – 1.08) | 1.01 (0.89 – 1.15) |
| **Subgroup <90 days (n)** | 67 | 61 | 68 | 69 |
| Attended clinic appointment, N (%) | 37 (55.2%) | 45 (73.8%) | 46 (67.7%) | 40 (58.0%) |
| Unadjusted risk ratio (95% CI) | 1 | 1.34 (1.02 – 1.74)** | 1.22 (0.93 – 1.61) | 1.05 (0.78 – 1.41) |
| Adjusted risk ratio* (95% CI) | 1 | 1.37 (1.09 – 1.75) | 1.19 (0.95 – 1.51) | 1.08 (0.85 – 1.39) |
| **90-181 days (n)** | 53 | 58 | 65 | 58 |
| Attended clinic appointment, N (%) | 33 (62.3%) | 35 (60.3%) | 31 (47.7%) | 32 (55.2%) |
| Unadjusted risk ratio (95% CI) | 1 | 0.96 (0.72 – 1.30) | 0.76 (0.55 - 1.07) | 0.89 (0.64 - 1.21) |
| Adjusted risk ratio* (95% CI) | 1 | 0.99 (0.76 – 1.28) | 0.84 (0.64 - 1.12) | 0.91 (0.69 - 1.22) |
| **182-365 days (n)** | 67 | 75 | 62 | 75 |
| Attended clinic appointment, N (%) | 38 (56.7%) | 43 (57.3%) | 30 (48.4%) | 48 (64.0%) |
| Unadjusted risk ratio (95% CI) | 1 | 1.01 (0.76 – 1.34) | 0.85 (0.61 - 1.19) | 1.12 (0.86 – 1.48) |
| Adjusted risk ratio* (95% CI) | 1 | 0.91 (0.71 - 1.18) | 0.79 (0.59 - 1.06) | 1.00 (0.78 – 1.28) |
| **>365 days (n)** | 193 | 195 | 181 | 187 |
| Attended clinic appointment, N (%) | 83 (43.0%) | 85 (43.6%) | 73 (40.3%) | 78 (41.7%) |
| Unadjusted risk ratio (95% CI) | 1 | 1.01 (0.81 - 1.27) | 0.93 (0.74 - 1.19) | 0.96 (0.77 - 1.23) |
| Adjusted risk ratio* (95% CI) | 1 | 1.01 (0.82 - 1.25) | 0.98 (0.78 - 1.24) | 1.01 (0.81 - 1.27) |

\* Adjusted for age, sex, clinic, purpose of visit, reminder call, and if visit was scheduled during month end. \*\* Evidence of differences in intervention effect size by duration on ART (unadjusted).

In the subgroup analysis, loss aversion framed messages increased the likelihood for timely clinic visits among participants who were on ART for <90 days (ARR 1.37; 95% CI: 1.08-1.74). Social norms messages and altruism-framed messages did not appear to be effective in any of the sub-groups that were examined.

## Discussion

Using behavioural science principles to frame text message reminders did not increase the likelihood of timely attendance of scheduled clinic visits among ART recipients in South Africa. Since appointment reminders are often sent in the form of text messages, we tested messages that drew upon loss aversion, social norms, and altruism to encourage care recipients to come to clinic for their appointment. In addition, the study also found that nearly 50% of care recipients, in all study arms, did not have timely clinic visits. This result underscores the continued need for alternative interventions that can promote clinic visits and retention in care more generally.

There are several reasons why the enhanced text messages may have been ineffective in promoting timely clinic attendance. First, nearly 40% of all text messages were not delivered to participants since many individuals routinely change their phone numbers, share numbers with others, or provide incorrect information to clinics (14,20,21). Verifying care recipients’ contact information at regular intervals is likely to be necessary for effective use of enhanced text messaging systems. Second, it could be that reasons for not having timely clinic attendance have been due to structural and economic barriers such as high transportation costs, opportunity costs of time, or long clinic wait times. These barriers have been shown to significantly impact clinic attendance, particularly for marginalized and underserved populations (22,23), and they are less likely to be addressed by text messages. Third, the lack of timely clinic attendance may be due to the perception of a scheduled clinic visit date as merely being a suggested date to come to the clinic rather than an exact appointment date. In some cases, there may be a mismatch between scheduled clinic visits and dispensation of medicines, particularly for those on treatment for longer periods.

Our subgroup analyses showed that among care recipients who had been on treatment for <90 days, loss aversion messages were effective in promoting timely clinic attendance. This is notable since research shows that care recipients who miss visits within the first six months on treatment have increased likelihood of poor outcomes (22,24). Care recipients who have been on treatment for >90 days may have become accustomed to clinic messages or may feel it is less important to come to the clinic on or before the scheduled appointment date especially if they still have medication in hand. Other studies have also shown a diminishing effect of messages over time or at various stages of treatment journey (17,25). It may be necessary to tailor messaging interventions to the specific stage of treatment to maximise their effectiveness.

### Study limitations

Our study has some limitations. The primary outcome was attending a scheduled visit on or before the appointment date, but we were unable to obtain information on whether participants came to the clinic after the scheduled appointment date. This may result in an underestimate of whether participants come to the clinic within a few days around the time of their appointment (before or after). However, it is less plausible that this affects the estimated effect sizes of the enhanced text messages. Second, we did not assess the impact of the interventions on long-term outcomes such as retention in care or viral suppression since our goal was to rapidly assess whether the messages changed behavior. Third, a large proportion of participants were scheduled to attend medication refill visits. This restricted us to understanding only one aspect of HIV service delivery and the effect of enhanced text messages. Lastly, we did not have a pure control that did not receive any text message, which may have prevented us from seeing an effect of any message compared to no message.

## Conclusions

Our study has implications on programmatic use of SMS reminders to improve clinic attendance. Behavioural science framed messages may be an effective, low-cost tool for increasing timely visit attendance among ART care recipients. However, the effectiveness of differently framed messages may depend on one’s stage of the treatment journey, and care recipients may be more responsive during the earlier stages. More work is needed to address the systematic and technological deficiencies that may reduce the impact of these interventions and to explore approaches to tailor these messages and apply them to the subgroups that would benefit the most.

## Data Availability

All data produced in the present study are available upon reasonable request to the authors

## Authors’ contributions

CG, CC, HT, SP, and TM designed the study. CG and TM had primary oversight of the study implementation. CG and TM conducted the analyses. CG prepared the first draft with input from HT. All authors reviewed and edited the draft.

## Acknowledgements

We would like to thank the all the participants and staff including but not limited to case officers, data capturers and nurses at the facilities we conducted the research for their support and contribution to the success of the study.

